# Discrimination of SARS-Cov 2 and arboviruses (DENV, ZIKV and CHIKV) clinical features using machine learning techniques: a fast and inexpensive clinical screening for countries simultaneously affected by both diseases

**DOI:** 10.1101/2021.01.28.21250714

**Authors:** João Daniel S. Castro

## Abstract

SARS-Cov-2 (Covid-19) has spread rapidly throughout the world, and especially in tropical countries already affected by outbreaks of arboviruses, such as Dengue, Zika and Chikungunya, and may lead these locations to a collapse of health systems. Thus, the present work aims to develop a methodology using a machine learning algorithm (Support Vector Machine) for the prediction and discrimination of patients affected by Covid-19 and arboviruses (DENV, ZIKV and CHIKV). Clinical data from 204 patients with both Covid-19 and arboviruses obtained from 23 scientific articles and 1 dataset were used. The developed model was able to predict 93.1% of Covid-19 cases and 82.1% of arbovirus cases, with an accuracy of 89.1% and Area under Roc Curve of 95.6%, proving to be effective in prediction and possible screening of these patients, especially those affected by Covid-19, allowing early isolation.

## Introduction

In December 2019, a series of pneumonia cases of unknown cause emerged among visitors to a wet market in the city of Wuhan, Hubei (China). Genetic sequencing of samples from the respiratory tract of these patients identified a new type of Coronavirus, called (2019-nCoV) [1], which since the outbreak, has affected more than 1 million people and totals 60,000 deaths worldwide [2,3].

Covid-19 produces a systemic respiratory disease that can progress to severe pneumonia in approximately 16% of cases [4]. Sepsis is the most frequent complication, followed by respiratory failure, acute respiratory distress (ARDS), heart failure and septic shock [5].

Concomitantly with the Covid-19 outbreak, tropical countries have faced successive outbreaks of arboviruses, such as Dengue (DENV), Zika (ZIKV) and Chikungunya (CHIKV), presenting themselves as a challenge to the health systems of these countries. Arboviruses are diseases caused by viruses transmitted to humans through the bites of infected blood-sucking arthropods, predominantly *Aedes* mosquitoes [6], and their clinical manifestations can vary from mild and undifferentiated febrile disease to neurological, joint and hemorrhagic febrile syndromes [7]. In addition, it is observed that there is a low probability of predicting a possible outbreak [8], which may surprise health systems that, today, are saturated due to the Covid-19 pandemic.

The imminence of a syndemic state between Covid-19 and arboviruses (mainly Dengue) has been shown to be a concern of researchers from Latin American countries [9,10]. Even though it has not yet reported cases of co-infection, your treatment may be adversely affected, since the spectrum of drugs does not seem compatible [11]. Associated with this, researchers have reported a false positive diagnosis for Dengue in situations in which patients are affected by the SARS-CoV-2 virus [12,13], even though there was no genomic similarity between the viruses [14], which can make the diagnosis of these diseases even more difficult.

Brazil has historically faced cycles of arbovirus epidemics throughout its territory, with Dengue being the most prevalent arbovirus [15]. According to the most recent Brazilian epidemiological bulletin, until now (ie, April 06, 2020) approximately 500 thousand cases of arboviruses have been reported (DENV, ZIKV, CHIKV), 96% of which are associated with dengue, 2.73% Chikungunya and 0.33% to Zika [16], compared to 9506 cases of Covid-19 (until April 3) [17].

Thus, in order to optimize resources and care for patients, as well as to enable effective screening, allowing isolation and early management, biomarkers (obtained through blood counts at admission) can be effective in predicting the degree of severity of disease. Studies indicate that Covid-19 causes Lymphocytopenia [4] and Thrombocytopenia [18], while arboviruses can cause Leukopenia, Lymphohistiocytosis and mild Lymphocytopenia [19–22].

Therefore, based on the clinical data of patients affected by Covid-19 or Arboviruses (DENV, ZIKV and CHIKV), this study aims to build a predictive model using machine learning algorithms, in order to screen and diagnose patients in case a possible syndemic state. It is also intended to share clinical data by the medical community about diseases of global circulation, in order to mitigate their impacts.

## 2 Materials and Methods

An electronic search was performed in the databases of Periódicos CAPES (portal of the Brazilian development agency CAPES, which aggregates numerous publishers of scientific journals), Google Scholar, Google Dataset and Science Direct, using the keywords, “Clinical features” OR “White blood cells count “OR” Haemogram “AND” coronavirus 2019 “OR” COVID-19 “OR” 2019-nCoV “OR” SARS-CoV-2 “. To search for clinical data related to arboviruses, the terms “Clinical features” OR “White blood cells count” OR “Haemogram” AND “Dengue” OR “Zika” OR “Chikungunya” were used. A Dataset containing clinical data from Brazilian patients Positive for SARS-CoV-2 was also added to the data set, made available by Hospital Israelita Albert Einstein [23].

The papers were chosen according to the criteria of: analysis of the abstract, title and body of the text, excluding works that presented only the average of the data obtained for a group of patients or that did not present the patients’ blood count. Thus, 23 scientific papers were selected, all of which were written in English.

This study was carried out in accordance with the Declaration of Helsinki and in accordance with the terms of local legislation.

The predictive model was built using the open-source software Orange (v. 3.20.1) [24]. Before the implementation of the algorithm, the data were normalized (center by mean) and then submitted to the detection of outliers (abnormal data), using covariance assessment.

The input data for the training of the algorithm were the White Blood Cells count (WBC) and the Lymphocite count, available in the articles, and the classes used for the prediction were Covid-19 and Arboviruses. The type of algorithm used was SVM with Cost regression (c) = 1.50, Kernel type RBF (g = 1,04) and the following optimization parameters: numerical tolerance = 0.005, without iteration limit.

The model was trained using the data and validated using the cross-validation methodology, with *k*-fold =20. The validation method consists of dividing the data set into *k* parts, using *k*-1 parts for modeling (training) and the remaining part for testing. In this model the data set was tested twenty times, each one with a different fold from dataset.

The idea behind this type of algorithm is to map the data, transposing it into a high dimensional space in which hyperplanes separate the data into subgroups (clusters) according to their characteristics, allowing the classification problem to be more easily solved [25].

The model developed is available in supplementary materials.

## 3. Results and discussion

### 3.1 Characteristics of the included studies

The detailed characteristics of this study are found in the Dataset available as supplementary material. Data from 204 patients were used in total, and after detecting outliers, 120 patients remained positive for SARS-CoV-2 and 67 patients with arboviruses (DENV, ZIKV and CHIKV), in a wide age group and of both sexes, totaling 187 patients.

The parameters used for the evaluation and screening was the existence / prevalence of Leukopenia and Lymphohistiocytosis / Lymphocytopenia, based on what was previously reported in the literature. Tables 1 and 2 summarize the data found in the literature. It is worth noting that the dataset provided by Hospital Israelita Albert Einstein [23] was already normalized, and therefore it is not shown in the data set in the tables below.

**Table 1.** Clinical features of patients affected by Covid-19

| Número de Pacientes | Disease | WBC ($10^9/L$) | Lymphocytes ($10^9/L$) | Observation | Reference |
| --- | --- | --- | --- | --- | --- |
| 9 | Covid-19 | 7.70 (5.07 -10.61) | 1.18 (0.46-1.59) | Pregnant | [26] |
| 5 | Covid-19 | 7.6 (5.03 - 10.76) | 0.74 (0.63 - 0.92) | Hemodialysis | [27] |
| 5 | Covid-19 | 8.75 (6.12 - 12,4) | 1.42 (0.84-2.46) | Asymptomatic | [28] |
| 5 | Covid-19 | 5.68 (4.4 - 7.1) | 1.56 (1.3- 1.7) | Reactivation | [29] |
| 6 | Covid-19 | 6.13 (4.3 – 11.4) | 1.7 (0.6 – 2.7) | - | [30] |
| 4 | Covid-19 | 5.49 (4.23 – 6.84) | 1.19 (0.42 - 1.98) | - | [31] |
| 2 | Covid-19 | 5.76 (4.37 - 7.16) | 1.03 (0.88 – 1.19) | - | [32] |
| 1 | Covid-19 | 10.6 | 0.86 | Pregnant | [33] |
| Total = 37 |  | Mean = 7,06 | Mean = 1,27 |  |  |

**Table 2.**
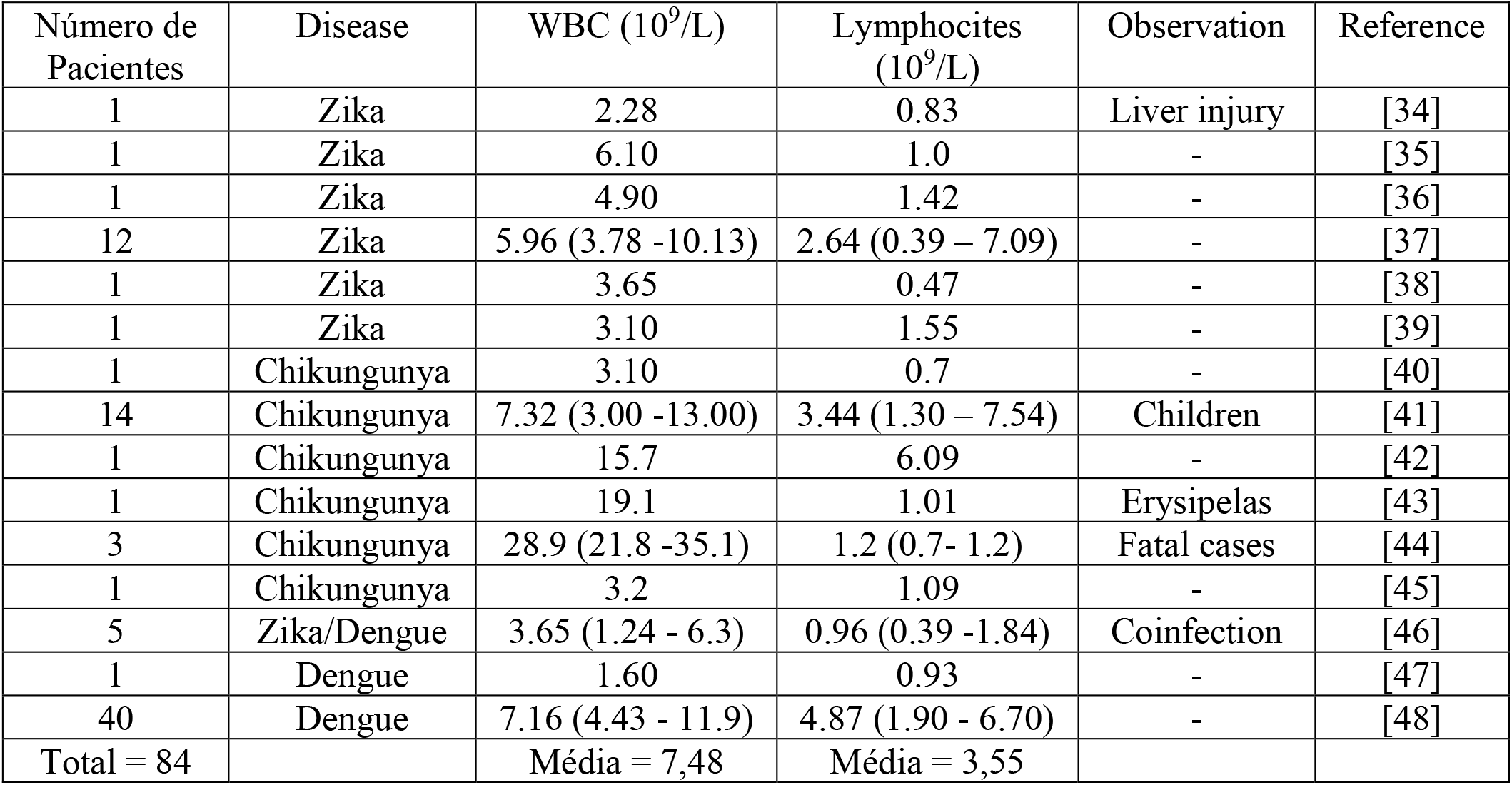
Clinical features of patients affected by arboviruses (DENV, ZIKV and CHIKV)

### 3.2 Model assessment

**Figure 1.**
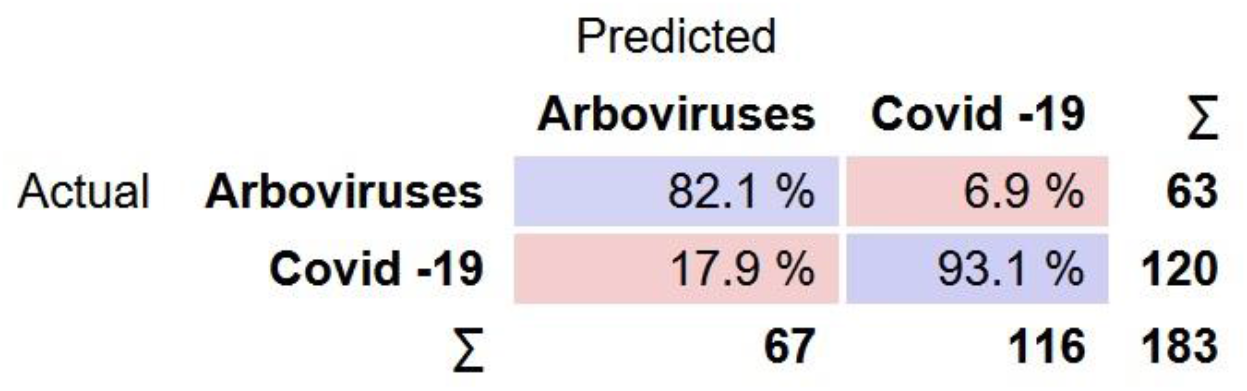
Confusion Matrix for classification test using an SVM machine learning algorithm

The results of a classification can be represented as a matrix called a confusion matrix, a square matrix (G x G) whose rows and columns represent the experimental and predicted data respectively [49].

The matrix shows that the model was able to predict more than 90% of Covid-19 cases and 82.1% of cases of arboviruses, based on clinical data obtained in the literature. The developed model also has an Area under Roc Curve (AUC) of 0.956, an accuracy of 0.891 and an precision of 0.893.

Lymphocytopenia is a clinical condition observed in patients with SARS-CoV-2, regardless of gender, age and / or pre-existence of comorbidities [50–52].

This condition allowed the correct classification of most of the observed cases. The classification error of individuals affected by arboviruses is also reported by Yan and collaborators, when describing two clinical pictures of SARS-Cov-2 as false positives for Dengue (DENV) [12], showing the urgency in the development of more accurate methodologies.

Leukopenia presents to patients with arboviruses (DENV, ZIKV and CHIKV) in medium to intense degrees, according to the PAHO (Pan-American Health Organization) classification [53], and also presented in clinical studies [54,55]. Associated with this, some patients may experience lymphohistiocytosis [56–58]. These characteristics allowed the correct classification of 82.1% of the cases.

Cases misclassified totaling 10.93% of cases and are presented in Table 3:

**Table 3.** Misclassified cases

| WBC ($10^9/L$) | Lymphocyte ($10^9/L$) | Actual | Predicted |
| --- | --- | --- | --- |
| 5.6 | 2.2 | Covid -19 | Arboviruses |
| 8.24 | 2.46 | Covid -19 | Arboviruses |
| 6.48 | 1.98 | Covid -19 | Arboviruses |
| 4.2 | 0.7 | Covid -19 | Arboviruses |
| 4.23 | 1.28 | Covid -19 | Arboviruses |
| 6.5 | 2.8 | Covid -19 | Arboviruses |
| 11.4 | 2.7 | Covid -19 | Arboviruses |
| 7.63 | 2.83 | Covid -19 | Arboviruses |
| 4.4 | 1.08 | Covid -19 | Arboviruses |
| 4.4 | 1.7 | Covid -19 | Arboviruses |
| 4.3 | 1.2 | Covid -19 | Arboviruses |
| 4.5 | 1.4 | Covid -19 | Arboviruses |
| 7.56 | 2.12 | Arboviruses | Covid -19 |
| 9.6 | 1.3 | Arboviruses | Covid -19 |
| 7.54 | 1.66 | Arboviruses | Covid -19 |
| 6.3 | 1.84 | Arboviruses | Covid -19 |
| 6.1 | 1 | Arboviruses | Covid -19 |
| 4.28 | 0.39 | Arboviruses | Covid -19 |
| 5.86 | 1.76 | Arboviruses | Covid -19 |
| 9.7 | 1.9 | Arboviruses | Covid -19 |

From the analysis of table 3 and figure 2, we can find the factors that allow the erroneous classification of patients: half of the patients affected by Covid-19 who have WBC values (≅ 5) associated with low values of lymphocytes (> 2) are classified as having arboviruses; while the other half is classified according to the high value of lymphocytes

Patients affected by arboviruses who have lymphocytopenia (lymphocytes ≤ 2) and without leukopenia are classified as having Covid-19.

**Figure 2.**
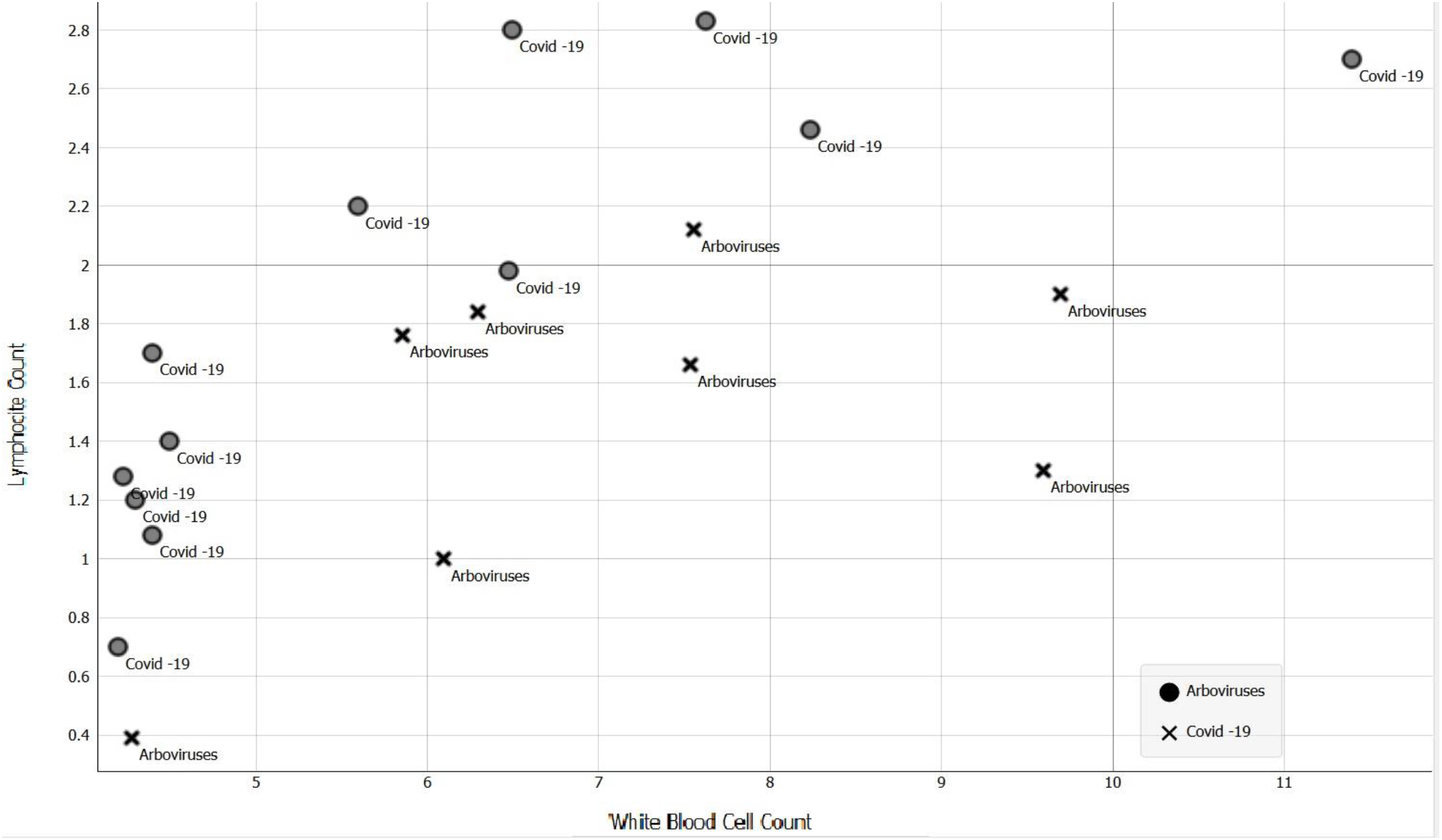
WBC x Lymphocite count plot for patients classified incorrectly. The caption of each patient indicates the disease to which it is affected, the symbols indicate the predicted illness

## 4 Conclusion

The model developed was effective in discriminating based on clinical data between patients affected by Covid-19 and patients affected by arboviruses (DENV, ZIKV, CHIKV), being able to predict 93.1% of the cases of SARS-CoV-2 and 82.1% of cases of arboviruses. Even though it presented a not very satisfactory result for arboviruses, the model allows the screening of patients with Coronavirus (contagious), thus speeding up the isolation and hospitalization procedures.

Because it was developed using free software, this model can be used in public health systems in countries in the event of a simultaneous outbreak of arboviruses and Covid-19, minimizing the impacts of a possible collapse of the health system.

## Data Availability

All data are available in a external dataset

https://data.mendeley.com/datasets/m9dcx3b5fm/1

## Acknowledgements

The author thanks the collaboration of doctors Marjorie Lobo and Tainá Cajazeira and the researcher Luane Barbosa for the review of this work.

## References

[1] C. Huang, Y. Wang, X. Li, L. Ren, J. Zhao, Y. Hu, L. Zhang, G. Fan, J. Xu, X. Gu, Clinical features of patients infected with 2019 novel coronavirus in Wuhan, China, Lancet. (2020) 497–506. doi:10.1016/S0140-6736(20)30183-5.

[2] World Health Organization, Coronavirus disease 2019 (COVID-19) Situation Report – 76, 2019 (2020). https://www.who.int/docs/default-source/coronaviruse/situation-reports/20200405-sitrep-76-covid-19.pdf?sfvrsn=6ecf0977_4.

[3] E. Dong, H. Du, L. Gardner, An interactive web-based dashboard to track COVID-19 in real time, Lancet Infect. Dis. 3099 (2020) 19–20. doi:10.1016/S1473-3099(20)30120-1.

[4] W. Guan, Z. Ni, Y. Hu, W. Liang, C. Ou, J. He, L. Liu, H. Shan, C. Lei, D.S.C. Hui, B. Du, L. Li, G. Zeng, K. Yuen, R. Chen, C. Tang, T. Wang, P. Chen, J. Xiang, S. Li, J. Wang, Z. Liang, Y. Peng, L. Wei, Y. Liu, Y. Hu, P. Peng, J. Wang, J. Liu, Z. Chen, G. Li, Z. Zheng, S. Qiu, J. Luo, C. Ye, S. Zhu, Clinical Characteristics of Coronavirus Disease 2019 in China, N. Engl. J. Med. (2020) 1– 13. doi:10.1056/NEJMoa2002032.

[5] F. Zhou, T. Yu, R. Du, G. Fan, Y. Liu, Z. Liu, J. Xiang, Y. Wang, B. Song, X. Gu, L. Guan, Y. Wei, Clinical course and risk factors for mortality of adult inpatients with COVID-19 in Wuhan, China?: a retrospective cohort study, Lancet. 395 (2020) 1054–1062. doi:10.1016/S0140-6736(20)30566-3.

[6] A. Wilder-smith, D.J. Gubler, S.C. Weaver, T.P. Monath, D.L. Heymann, T.W. Scott, Epidemic arboviral diseases?: priorities for research and public health, Lancet Infect. Dis. 3099 (2016) 1–6. doi:10.1016/S1473-3099(16)30518-7.

[7] A.P.B. Von Zuben, Maria Rita Donalisio, André Ricardo Ribas Freitas, Arboviroses emergentes no Brasil?: desafios para a clínica e implicações para a saúde pública, Rev. Saude Publica. 51 (2017) 10–15.

[8] D. Musso, A.J. Rodriguez-morales, J.E. Levi, D.J. Gubler, Personal View Unexpected outbreaks of arbovirus infections?: lessons learned from the Pacific and tropical America, Lancet Infect. Dis. 3099 (2018) 1–7. doi:10.1016/S1473-3099(18)30269-X.

[9] A.J. Rodriguez-Morales, V. Gallego, J.P. Escalera-Antezana, C.A. Méndez, L.I. Zambrano, C. Franco-Paredes, J.A. Suárez, H.D. Rodriguez-Enciso, G.J. Balbin-Ramon, E. Savio-Larriera, A. Risquez, S. Cimerman, COVID-19 in Latin America: The implications of the first confirmed case in Brazil, Travel Med. Infect. Dis. (2020) 101613. doi:10.1016/j.tmaid.2020.101613.

[10] M. Saavedra-Velasco, C. Chiara-Chilet, R. Pichardo-Rodriguez, A. Grandez-Urbina, F. Inga-Berrospi, Coinfección entre dengue y COVID-19: Necesidad de abordaje en zonas endémicas, Rev. Fac. Cienc. Med. Cordoba. 77 (2020) 52–54. doi:10.31053/1853.0605.v77.n1.28031.

[11] J. Navarro, J. Arrivillaga-henríquez, J. Salazar-loor, A.J. Rodriguez-morales, COVID-19 and dengue, co-epidemics in Ecuador and other countries in Latin America: Pushing strained health care systems over the edge, Travel Med. Infect. Dis. (2020) 101656. doi:10.1016/j.tmaid.2020.101656.

[12] G. Yan, C.K. Lee, L.T.M. Lam, B. Yan, Y.X. Chua, A.Y.N. Lim, K.F. Phang, G. Sen Kew, H. Teng, C.H. Ngai, L. Lin, R.M. Foo, S. Pada, L.C. Ng, P.A. Tambyah, Covert COVID-19 and false-positive dengue serology in Singapore, Lancet Infect. Dis. 3099 (2020) 30158. doi:10.1016/S1473-3099(20)30158-4.

[13] B. joob, V. Wiwanitkit, COVID-19 can present with a rash and be mistaken for Dengue, J. Am. Acad. Dermatol. (2020) 19–22. doi:10.1016/j.jaad.2020.03.036.

[14] Y. Li, B. Liu, J. Cui, Z. Wang, Y. Shen, Y. Xu, K. Yao, Y. Guan, Similarities and Evolutionary Relationships of COVID-19 and Related Viruses, (2020). http://arxiv.org/abs/2003.05580.

[15] V.I. Avelino-Silva, J.F. Ramos, Arboviroses e políticas públicas no Brasil Arboviruses and public policies in Brazil, Rev. Ciências Em Saúde. 7 (2017) 1–2. doi:10.21876/rcsfmit.v7i3.675.

[16] Brasil, Monitoramento dos casos de arboviroses urbanas transmitidas pelo Aedes Aegypti (dengue, chikungunya e zika), Semanas Epidemiológicas 1 a 13, 2020, Bol. Epidemiológico 14. 51 (2020) 1–34. https://www.saude.gov.br/images/pdf/2020/April/03/Boletim-epidemiologico-SVS-14.pdf.

[17] Brasil, Doença pelo Coronavírus 2019, Bol. Epidemiológico Do Cent. Operações Emergência Em Saúde Pública 06. 06 (2020) 1–23. https://portalarquivos.saude.gov.br/images/pdf/2020/April/03/BE6-Boletim-Especial-do-COE.pdf.

[18] G. Lippi, M. Plebani, B. Michael, Thrombocytopenia is associated with severe coronavirus disease 2019 (COVID-19) infections?: A meta-analysis, Clin. Chim. Acta. 506 (2020) 145–148. doi:10.1016/j.cca.2020.03.022.

[19] T. Elias, C. Fernanda, A. Francesli, N. Reis, N. Santos, M. Lima, E. Márcia, S. Cabrera, I. Nuremberg, F. Rodrigues, L. Elisa, A. Antônio, Clinical, laboratory and virological data from suspected ZIKV patients in an endemic arbovirus area, J. Clin. Virol. 96 (2017) 20–25. doi:10.1016/j.jcv.2017.09.002.

[20] M. Kapdi, I.R.A. Shah, Dengue and haemophagocytic lymphohistiocytosis, Scand. J. Infect. Dis. (2012) 708–709. doi:10.3109/00365548.2011.652667.

[21] S.-C.J. Wæhre T, Maagard A, Tappe D, Cadar D, Zika Virus Infection after Travel to Tahiti, December 2013, Emerg. Infect. Dis. 20 (2014) 8–10.

[22] L. Dupuis-Maguiraga, M. Noret, S. Brun, R. Le Grand, G. Gras, P. Roques, Chikungunya Disease?: Infection-Associated Markers from the Acute to the Chronic Phase of Arbovirus-Induced Arthralgia, PLoS Negl. Trop. Dis. 6 (2012). doi:10.1371/journal.pntd.0001446.

[23] E. Data4u, Diagnosis of COVID-19 and its clinical spectrum, Kaggle. (2020). https://www.kaggle.com/einsteindata4u/covid19 (accessed April 5, 2020).

[24] J. Demsar, B. Zupan, G. Leban, T. Curk, Orange?: From Experimental Machine Learning, Knowl. Discov. Databases PKDD 2004. (2004) 537–539. doi:10.1007/978-3-540-30116-5_58.

[25] J. Luts, F. Ojeda, R. Van de Plas Raf, B. De Moor, S. Van Huffel, J.A.K. Suykens, A tutorial on support vector machine-based methods for classification problems in chemometrics, Anal. Chim. Acta. 665 (2010) 129–145. doi:10.1016/j.aca.2010.03.030.

[26] H. Chen, J. Guo, C. Wang, F. Luo, X. Yu, W. Zhang, J. Li, D. Zhao, D. Xu, Q. Gong, J. Liao, H. Yang, W. Hou, Y. Zhang, Clinical characteristics and intrauterine vertical transmission potential of COVID-19 infection in nine pregnant women: a retrospective review of medical records, Lancet. 395 (2020) 809–815. doi:10.1016/S0140-6736(20)30360-3.

[27] R. Wang, C. Liao, H. He, C. Hu, Z. Wei, Z. Hong, C. Zhang, M. Liao, H. Shui, COVID-19 in Hemodialysis Patients: A Report of 5 Cases, Am. J. Kidney Dis. (2020). doi:10.1053/j.ajkd.2020.03.009.

[28] F. Ye, S. Xu, Z. Rong, R. Xu, X. Liu, P. Deng, H. Liu, X. Xu, Delivery of infection from asymptomatic carriers of COVID-19 in a familial cluster, Int. J. Infect. Dis. 105 (2020) 72–80. doi:10.1016/j.ijid.2020.03.042.

[29] G. Ye, Z. Pan, Y. Pan, Q. Deng, L. Chen, J. Li, Y. Li, X. Wang, Clinical characteristics of severe acute respiratory syndrome coronavirus 2 reactivation, J. Infect. (2020) 3–6. doi:10.1016/j.jinf.2020.03.001.

[30] J.F.W. Chan, S. Yuan, K.H. Kok, K.K.W. To, H. Chu, J. Yang, F. Xing, J. Liu, C.C.Y. Yip, R.W.S. Poon, H.W. Tsoi, S.K.F. Lo, K.H. Chan, V.K.M. Poon, W.M. Chan, J.D. Ip, J.P. Cai, V.C.C. Cheng, H. Chen, C.K.M. Hui, K.Y. Yuen, A familial cluster of pneumonia associated with the 2019 novel coronavirus indicating person-to-person transmission: a study of a family cluster, Lancet. 395 (2020) 514–523. doi:10.1016/S0140-6736(20)30154-9.

[31] Z. Wang, X. Chen, Y. Lu, F. Chen, W. Zhang, Clinical characteristics and therapeutic procedure for four cases with 2019 novel coronavirus pneumonia receiving combined Chinese and Western medicine treatment, Biosci. Trends. 14 (2020) 64–68. doi:10.5582/bst.2020.01030.

[32] P. Yu, J. Zhu, Z. Zhang, Y. Han, L. Huang, A familial cluster of infection associated with the 2019 novel coronavirus indicating potential person-to-person transmission during the incubation period, J. Infect. Dis. (2020) 1–5.doi:10.1093/infdis/jiaa077.

[33] X. Wang, Z. Zhou, J. Zhang, F. Zhu, Y. Tang, X. Shen, A case of 2019 Novel Coronavirus in a pregnant woman with preterm delivery, Clin. Infect. Dis. (2020). doi:10.1093/cid/ciaa200.

[34] Y. Wu, X. Cui, N. Wu, R. Song, W. Yang, W. Zhang, D. Fan, Z. Chen, J. An, A unique case of human Zika virus infection in association with severe liver injury and coagulation disorders, Sci. Rep. 7 (2017) 1–8. doi:10.1038/s41598-017-11568-4.

[35] G.H.Y. Leung, R.W. Baird, J. Druce, N.M. Anstey, Zika virus infection in Australia following a monkey bite in Indonesia, Southeast Asian J. Trop. Med. Public Health. 46 (2015) 460–464.

[36] H.C. Jang, W.B. Park, U.J. Kim, J.Y. Chun, S.J. Choi, P.G. Choe, S.I. Jung, Y. Jee, N.J. Kim, E.H. Choi, M.D. Oh, First imported case of Zika virus infection into Korea, J. Korean Med. Sci. 31 (2016) 1173–1177. doi:10.3346/jkms.2016.31.7.1173.

[37] J. Li, C.Y. Chong, N.W. Tan, C.F. Yung, N.W. Tee, K.C. Thoon, Characteristics of Zika Virus Disease in Children: Clinical, Hematological, and Virological Findings from an Outbreak in Singapore, Clin. Infect. Dis. 64 (2017) 1445–1448. doi:10.1093/cid/cix137.

[38] L.H. Chen, Zika Virus Infection in a Massachusetts Resident After Travel to Costa Rica: A Case Report, Ann. Intern. Med. 164 (2016) 574. doi:10.7326/L16-0075.

[39] A.F. Cardona-Cardona, A.J. Rodríguez Morales, Severe abdominal pain in a patient with Zika infection: A case in Risaralda, Colombia, J. Infect. Public Health. 9 (2016) 372–373. doi:10.1016/j.jiph.2016.03.001.

[40] C. Therrien, G. Jourdan, K. Holloway, C. Tremblay, M.A. Drebot, First Imported Case of Chikungunya Virus Infection in a Travelling Canadian Returning from the Caribbean, Can. J. Infect. Dis. Med. Microbiol. 2016 (2016). doi:10.1155/2016/2980297.

[41] G.M. Oliveira, T.M.A. Marques, J.R. Dos Santos, E.D.F. Daher, R.D. Leite, E.S. Girão, R. da J.P. Neto, Clinical and laboratory profiles of children with sever chikungunya infection, Rev. Soc. Bras. Med. Trop. 52 (2019) 2017–2020. doi:10.1590/0037-8682-0232-2018.

[42] M.M. Aly, S. Ali, A.F. Muianga, V. Monteiro, J.G. Gallego, J. Weyer, K.I. Falk, J.T. Paweska, J. Cliff, E.S. Gudo, Severe Chikungunya infection in Northern Mozambique: a case report, BMC Res. Notes. 10 (2017) 4–9. doi:10.1186/s13104-017-2417-z.

[43] M.C. Schechter, K.A. Workowski, J.K. Fairley, Unusual Presentation of Chikungunya Virus Infection With Concomintant Erysipelas in a Returning Traveler From the Caribbean: A Case Report, Open Forum Infect. Dis. 1 (2014) ofu097–ofu097. doi:10.1093/ofid/ofu097.

[44] J.M. de la Hoz, B. Bayona, S. Viloria, J.L. Accini, H.S. Juan-Vergara, D. Viasus, Fatal cases of Chikungunya virus infection in Colombia: Diagnostic and treatment challenges, J. Clin. Virol. 69 (2015) 27–29. doi:10.1016/j.jcv.2015.05.021.

[45] I. Eckerle, V.T. Briciu, Ergönül, M. Lupşe, A. Papa, A. Radulescu, S. Tsiodras, C. Tsitou, C. Drosten, V.R. Nussenblatt, C.B. Reusken, L.A. Sigfrid, N.J. Beeching, Emerging souvenirs—clinical presentation of the returning traveller with imported arbovirus infections in Europe, Clin. Microbiol. Infect. 24 (2018) 240–245. doi:10.1016/j.cmi.2018.01.007.

[46] P.Y. Chia, H. Sen Yew, H. Ho, A. Chow, S.P. Sadarangani, M. Chan, Y.W. Kam, C.Y. Chong, K.C. Thoon, C.F. Yung, J.H. Li, D.C. Lye, P.P. De, L.F.P. Ng, T.W. Yeo, Y.S. Leo, Clinical features of patients with Zika and dengue virus co-infection in Singapore, J. Infect. 74 (2017) 611–615. doi:10.1016/j.jinf.2017.03.007.

[47] M. Mardani, F. Abbasi, M. Aghahasani, B. Ghavam, First Iranian imported case of dengue, Int. J. Prev. Med. 4 (2013) 1075–1077.

[48] S. Advani, S. Agarwal, J. Verma, Haemogram profile of dengue fever in adults during 19 September – 12 November 2008: A study of 40 cases from Delhi., Dengue Bull. 35 (2011) 71–75. https://apps.who.int/iris/handle/10665/171014.

[49] D. Ballabio, F. Grisoni, R. Todeschini, Multivariate comparison of classification performance measures, Chemom. Intell. Lab. Syst. 174 (2018) 33–44. doi:10.1016/j.chemolab.2017.12.004.

[50] L. Zeng, S. Xia, W. Yuan, K. Yan, F. Xiao, J. Shao, W. Zhou, Neonatal Early-Onset Infection with SARS-CoV-2 in 33 Neonates Born to Mothers with COVID-19 in Wuhan, China, JAMA Pediatr. 23 (2020) 4–6. doi:10.1001/jamapediatrics.2020.0878.

[51] X. Yang, Y. Yu, J. Xu, H. Shu, J. Xia, H. Liu, Y. Wu, L. Zhang, Z. Yu, M. Fang, T. Yu, Y. Wang, S. Pan, X. Zou, S. Yuan, Y. Shang, Clinical course and outcomes of critically ill patients with SARS-CoV-2 pneumonia in Wuhan, China: a single-centered, retrospective, observational study, Lancet Respir. Med. 2600 (2020) 1–7. doi:10.1016/S2213-2600(20)30079-5.

[52] D. Wu, T. Wu, Q. Liu, Z. Yang, The SARS-CoV-2 outbreak: what we know., Int. J. Infect. Dis. (2020). doi:10.1016/j.ijid.2020.03.004.

[53] Pan American Health Organization (PAHO), Tool for the diagnosis and care of patients with suspected arboviral diseases, 2017. http://iris.paho.org/xmlui/handle/123456789/33895.

[54] E.L. Azeredo, F.B. dos Santos, L.S. Barbosa, T.M.A. Souza, J. Badolato-Corrêa, J.C. Sánchez-Arcila, P.C.G. Nunes, L.M. de-Oliveira-Pinto, A.M. de Filippis, M. Dal Fabbro, I. Hoscher Romanholi, R. Cunha, Clinical and Laboratory Profile of Zika and Dengue Infected Patients: Lessons Learned From the Co-circulation of Dengue, Zika and Chikungunya in Brazil, PLoS Curr. (2018) 1–20. doi:10.1371/currents.outbreaks.0bf6aeb4d30824de63c4d5d745b217f5.

[55] R. Li, J. Ding, G. Ding, X. Fan, Y. He, X. Wang, H. Zhang, J. Ji, H. Li, Zika virus infections, a review, Radiol. Infect. Dis. 4 (2017) 88–93. doi:10.1016/j.jrid.2017.01.002.

[56] S.L. Beltrán-Silva, S.S. Chacón-Hernández, E. Moreno-Palacios, J.Á. Pereyra-Molina, Clinical and differential diagnosis: Dengue, chikungunya and Zika, Rev. Médica Del Hosp. Gen. México. 81 (2018) 146–153. doi:10.1016/j.hgmx.2016.09.011.

[57] E.M. Ellis, T.M. Sharp, J. Pérez-Padilla, L. González, B.K. Poole-Smith, E. Lebo, C. Baker, M.J. Delorey, B. Torres-Velasquez, E. Ochoa, B. Rivera-Garcia, H. Díaz-Pinto, L. Clavell, A. Puig-Ramos, G.E. Janka, K.M. Tomashek, Incidence and Risk Factors for Developing Dengue-Associated Hemophagocytic Lymphohistiocytosis in Puerto Rico, 2008 - 2013, PLoS Negl. Trop. Dis. 10 (2016) 2008–2013. doi:10.1371/journal.pntd.0004939.

[58] T.M. Sharp, L. Gaul, A. Muehlenbachs, E. Hunsperger, J. Bhatnagar, R. Lueptow, G.A. Santiago, J.L. Muñoz-Jordan, D.M. Blau, P. Ettestad, J.D. Bissett, S.C. Ledet, S.R. Zaki, K.M. Tomashek, Fatal hemophagocytic lymphohistiocytosis associated with locally acquired dengue virus infection - New Mexico and Texas, 2012, Morb. Mortal. Wkly. Rep. 63 (2014) 49–54. doi:10.1016/j.annemergmed.2014.04.007.

